# A Computational Framework for Identifying Age Risks in Drug-Adverse Event Pairs

**DOI:** 10.1101/2022.01.07.22268907

**Authors:** Zhizhen Zhao, Ruoqi Liu, Lei Wang, Lang Li, Chi Song, Ping Zhang

## Abstract

The identification of associations between drugs and adverse drug events (ADEs) is crucial for drug safety surveillance. An increasing number of studies have revealed that children and seniors are susceptible to ADEs at the population level. However, the comprehensive explorations of age risks in drug-ADE pairs are still limited. The FDA Adverse Event Reporting System (FAERS) provides individual case reports, which can be used for quantifying different age risks. In this study, we developed a statistical computational framework to detect age group of patients who are susceptible to some ADEs after taking specific drugs. We adopted different Chi-squared tests and conducted disproportionality analysis to detect drug-ADE pairs with age differences. We analyzed 4,580,113 drug-ADE pairs in FAERS (2004 to 2018Q3) and identified 2,523 pairs with the highest age risk. Furthermore, we conducted a case study on statin-induced ADE in children and youth. The code and results are available at https://github.com/Zhizhen-Zhao/Age-Risk-Identification

## Introduction

Adverse Drug Events (ADEs) are considered any noxious, unintended or undesired effect of a drug that occurs at a dose normally used in humans for prophylaxis, diagnosis or treatment^1^. ADEs are the 4th leading cause of death in the United States, which significantly increases the economic burden, stay length and death risk for hospitalized patients^2^. It is estimated that hospitalized patients have more than 2,216,000 ADEs in U.S., which cause more than 106,000 deaths annually^3^.

Age is an important risk factor to experience ADEs. Previous studies illustrated significant ADE differences among various age groups^4^. The 20-29 years old group has the lowest rate of adverse events, whereas the 0-9 years old group has the highest rate^5,6^. Young children have greater potential to report adverse events than adults, especially in the anti-infective, respiratory, dermatological and nervous system, because the detoxification mechanisms of children are immature^7^. Among children, a clinical trial showed that treating asthma through inhaled corticosteroids, younger children developed more cough and perioral dermatitis, while older children report hoarseness more frequently^5^. Furthermore, multiple studies have shown that the risk of an ADE increases with age^5,8^. For instance, with patent foramen ovale, older cryptogenic stroke patients have greater risks of having adverse events than younger patients^8^. Among patients with heart failure, risks of ADEs after taking digoxin enhance significantly with age, from 1.7% for patients below 50 to 5.4% for patients older than 80-year-old^5^. Although previous studies revealed certain ADEs with age risks, there still lacks comprehensive exploration, especially for ADEs associated with particular drugs.

To detect an association among interested drug-ADE pairs, data mining methods have been developed on measures of disproportionality, such as Reporting Odds Ratio (ROR) and the Proportional Reporting Ratio (PRR)^9^. However, many of them ignored heterogeneous characteristics of patients (e.g. age, gender and primary diseases) and gave the same weight to the information from the whole population when calculating the expected number of reports for a specific drug-ADE pair, which might mask the true signals or flag false associations as potential signals^12^.

The US Food and Drug Administration (FDA) provides post-marketing drug surveillance data Adverse Event Reporting System (FAERS)^13^, which collects data for suspected adverse drug events for further analysis. It covers a wide range of products aimed at diverse medical indications and are used across a broad range of patient populations. We aim to analyze the FAERS database and identify the age of patients who are more likely to experience certain ADEs after taking a specific drug. In precision medicine, a few studies adopted data-driven statistical subgroup methods to estimate average treatment effects and explore subgroups with enhanced treatment effects^10,11^. However, due to the lack of a control group, popular subgroup methods could not be applied in the database directly. Current researchers have used subgroup disproportionality to quantify the differential risk of a drug causing an ADE in men or women^14^ and have predicted the probability of being a female given confounding factors and built balanced cohorts to dampen the confounding biases existing in FAERS in the meantime^2^. But existing work is designed for detecting gender differences and examine ADEs in only two subpopulations (i.e., male and female). Their methods could not be directly leveraged for identifying ADEs in multiple age groups (i.e., children, youth, adult and senior).

In this study, we proposed a computational framework to identify the age group of patients who tend to experience more ADEs after taking a specific drug. Patients were divided into four age groups based on the World Population Prospects from The United Nations Department of Economic and Social Affairs^15^. Firstly, we performed Chi-squared tests to identify drug-ADE combinations which showed significant age differences. Then, for each drug-ADE pair, we conducted pairwise comparisons and measured disproportionality to detect the age group of patients with the highest risk of experiencing an adverse event. Finally, we applied logistic regression and likelihood ratio test (LRT) on the interaction between age group and drug to remove the confounding effects caused by age bias. We applied our framework on submissions of FAERS from 2004 to the third quarter of 2018. We successfully discovered 2,523 age-related drug-ADE combinations and identified the highest risk age group in each combination. We also conducted a case study on statin drugs induced ADE in children and youth.

Overall, our contributions can be summarized as follows:

- We developed a new computational framework to quantify the risks of experiencing ADEs for patients in multiple age subgroups. Our framework allows for the identification of heterogeneous characteristics of patients that have multiple categories. Through this computational framework, we fully explored age risks in drug-ADE pairs.
- To discover the highest risk among multiple age groups, we determined exactly which subgroups are significantly different in reporting an ADE, then calculated the reporting odds ratios (RORs) and selected the subgroup with the highest ROR as the significant signal.
- We applied our framework to discover age risks in drug-ADE pairs in FAERS data set. We also analyzed statin-related ADEs that have the highest risk in children and youth.

## Methodology

In our study, the age risks were detected mainly through four steps: discover age differences for specific drugs, discover age differences for specific drug-ADE pairs, discover the age group with higher risk, remove confounding effect by age bias. The process is presented in Algorithm 1. The illustration of our framework can be found in Figure 1.

**Figure 1:**
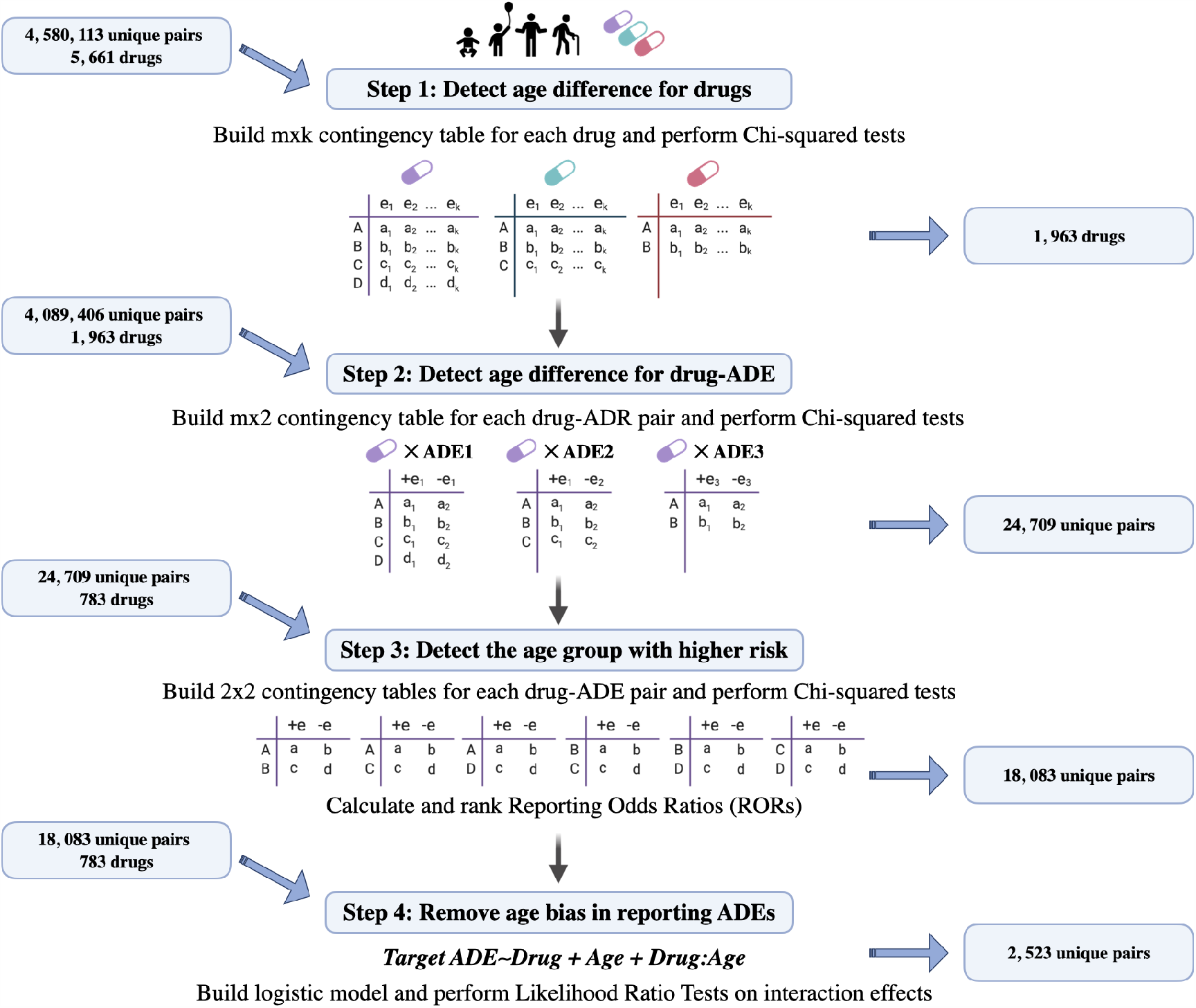
Illustration of the proposed methodology: In the first step, overall Chi-squared tests for each drug are performed to identify drugs with overall age differences. Then overall Chi-squared tests for each identified drugs and ADEs pair are performed to identify drug-ADE pairs with age differences. Next Chi-squared tests for age group comparisons within each pair are performed. RORs for every two age groups are computed and ranked, which quantifies the age risks. At the end, a logistic model is built for each detected pair and the Likelihood Ratio Test is performed on the interaction of drug and age group to remove age bias.

### Detect age differences for drugs

First, we detected age differences for a specific drug by considering all adverse drug events appearing with the drug. The goal was to discover drugs that have an overall age difference in drug-ADE pairs frequency distribution. An overall Chi-square test was applied to identify if there is an overall shift in drug-ADE pairs frequency distribution. Because of the large number of drug-ADE pairs, this step could also help avoid testing all individual unique pairs and then alleviate the testing burden. In addition, there were some drugs that only appeared in one particular age group, which we are not interested in. Thus, we filtered out drugs that appear only in one group and conducted tests on drugs that appear in more than two age groups.

For each drug, assume there were *k* ADEs in total that appeared with the drug. We constructed one of the contingency tables shown in Figure 1 based on how many age groups the drug appeared in. The value in each cell represents the total count of reports for each drug and the corresponding *ADE*_*i*_ (*i* = 1, …, *k*) in each age group.

We applied a Chi-square test for the contingency table, where the null hypothesis is that there is no age difference for a specific drug. For instance, for the drug occurring in all four groups, 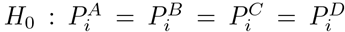, for all *i* = 1, …, *k*, where 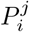 denotes the probability of being in the age group *j* and having the *ADE*_*i*_, capital letters represent four different age groups. Bonferroni methods were used for all drugs to adjust P values to correct for multiple testing.

#### Algorithm 1: The algorithmic framework to identify drug-ADE pairs with age risk

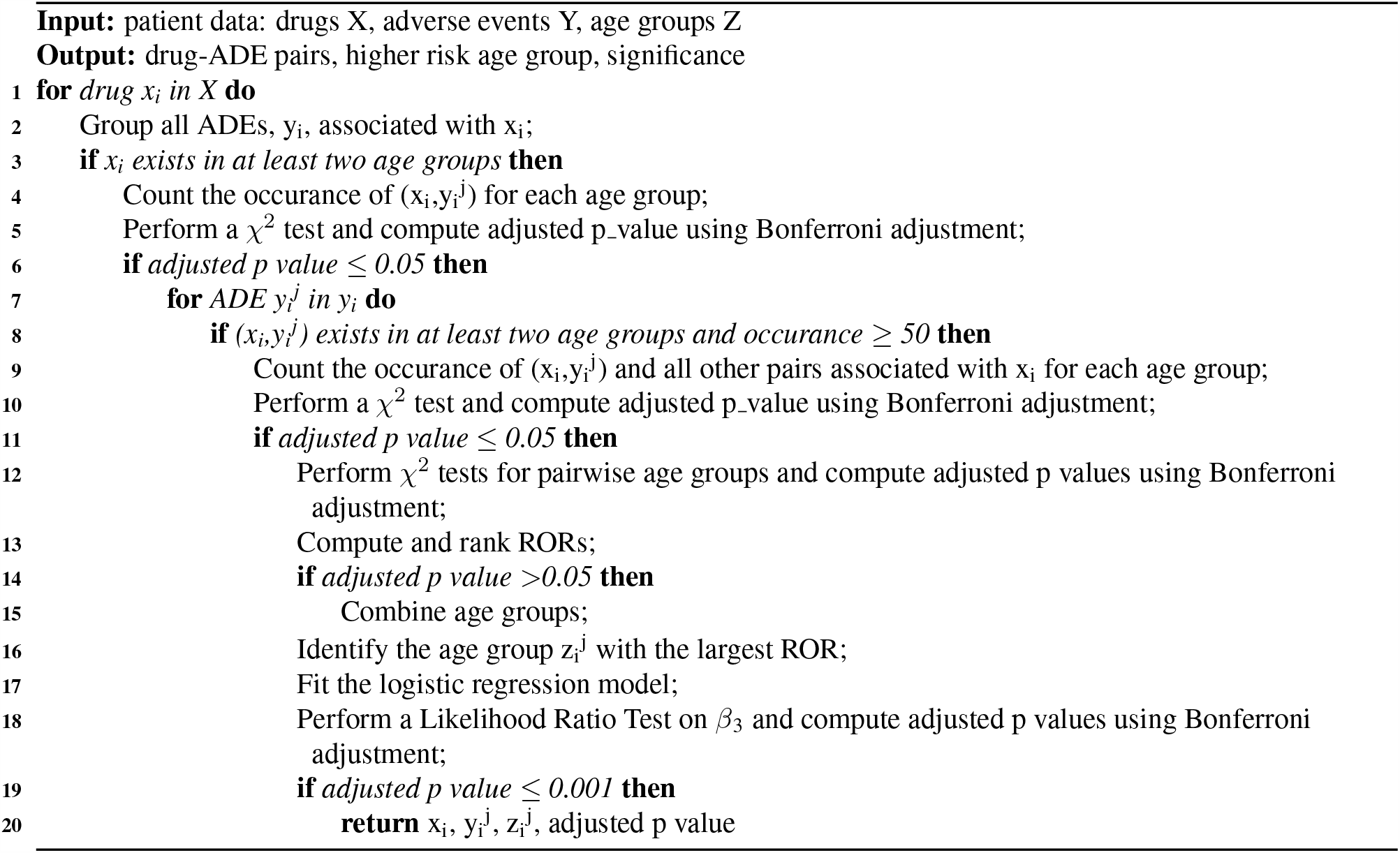

### Detect age differences for drug-ADE pairs

The first step filtered out the drugs that illustrated an overall difference in drug-ADE pairs frequency distribution, but not necessarily indicated that all pairs related to those drugs showed age differences. Therefore, this step is to apply a Chi-square test to identify if there exist age differences in a particular drug and adverse drug event pair. We are still only interested in the pairs that occurred in more than two age groups, so filtered out pairs that occurred only in one age group. In addition, in order to ensure the validity of Chi-square tests, we filtered out pairs whose occurrence was less than 50.

For each unique drug-ADE pair, we constructed one of the contingency tables shown in Figure 1 based on how many age groups the pair appeared in. The first column represents the number of reports for the interested adverse drug event after taking the interested drug for each age group. The second column represents the number of reports for all adverse drug events that occurred with the interested drug except the interested one for each age group. Based on the contingency table, we tested the null hypothesis that there is no age difference for a specific drug-ADE pair. The P values were adjusted through Bonferroni methods for all testing pairs to correct for multiple testing.

### Detect the age group with higher risk

The above two steps provided drug-ADE pairs that showed different distributions for various age groups, but they did not point out which groups accounted for the differences. We would then conduct pairwise comparisons and make disproportionality analyses for subpopulations of drug-ADE pairs obtained from the last step. The objective is to identify one or more age groups that are different from other age groups and are more likely to experience the target adverse event for a specific drug-ADE pair.

Since the pairs might exist in two, three or four age groups, we conducted at least one and at most six pairwise comparisons. Chi-square tests were conducted by constructing at least one and at most six 2* 2 contingency tables for unique drug-ADE pairs. For instance, if a pair existed in all four age groups, then six 2* 2 contingency tables shown in Figure 1 would be constructed. In order to ensure the validity of Chi-Square tests, all cells in all contingency tables are greater than five. Therefore, the drug-ADE pairs with occurrence more than zero, and less than five in at least one age group were filtered.

Meanwhile, adjusted ROR for each table was calculated. Adjusted ROR was defined as: 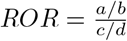, where a denotes the number of patients in the first age group experiencing the target ADE, b denotes the number of patients in the first age group experiencing all other ADEs except the target ADE, c denotes the number of patients in the second age group experiencing the target ADE, d denotes the number of patients in the second age group experiencing all other ADEs except the target ADE. A ROR greater than 1 indicated that the first age group is more likely to report the target ADE after taking the specific drug. In contrast, a ROR less than 1 indicated that the second age group is at a higher risk when experiencing the target ADE.

Risks for different age groups can be ordered through the value of ROR. Considering one group as a baseline and fix it, then RORs calculated in terms of that group can be ordered from the least value to the greatest value. On the basis of pairwise comparisons, we then combined the groups which are not significantly different. For groups that were not significantly different, to be more conservative, kept the ROR with the smallest value in the rank. Selected the age groups which have the highest risk as to the significant signals, meaning that certain age groups are susceptible to certain ADEs after taking a drug. For instance, all four group patients taking lamotrigine once experienced convulsion. There did not show a significant difference between children and youth but showed significant differences among the other five comparisons. Combining children group with youth group, and ordered the RORs, children and youth group illustrated the largest risk with the highest odd ratio. Thus we considered children and youth as higher risk groups, meaning that they tend to report more convulsion among the whole population.

In pairwise comparisons, we are only interested in pairs that showed consistent significantly different results within all comparisons. To be more specific, for a specific drug-ADE pair, if group A and group B are significantly different, group A and group C are significantly different, but group B and group C are not significantly different, we wouldn’t continue to analyze it. P values within each drug-ADE pair were adjusted through Bonferroni method for correction.

### Remove age bias in reporting ADEs

It’s possible that ADEs selected above are caused by age, not necessarily the interested drug. For instance, older adults are more likely to report inadequate pain management than younger adults. The previous Chi-square test might still identify the pairs associated with certain outcomes with the age difference, but these pairs may be not drug-specific. So we would filter out the potential confounding effects in reporting ADEs due to age bias. In other words, we are interested in discovering the ADEs that are caused by the interaction effects of drug and age. Thus, we conducted a logistic regression for each drug-ADE pair identified in the last step and tested on the interaction term. We build such a model:

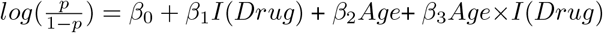

where Drug is an indicator variable, taking values on 1 if taking the target drug and 0 if not. Target ADE is 1 if experiencing the target ADE and 0 if experiencing other ADEs. *β*_0_ represents the log-odds of reporting the target ADE that the patient not taking the drug and in the first age group. *β*_1_ + *β*_3_ represents the increase of log-odds if taking the drug than not. *β*_3_ is the parameter that we are interested in, which quantifies the drug-age interaction effects. Thus, we conducted a likelihood ratio test on *β*_3_. The significant results illustrated that the drug effects depend on the identified high-risk age group. Bonferroni adjustment for all candidate drug-ADE pairs was used for multiple testings.

### Experiment

#### Data Set

In the source FAERS database, there exist multiple versions for an individual report, including one or more follow-up case versions based on the initial case version. In addition, the drug names in FAERS are not normalized, full names, trade names, abbreviations and spelling mistakes are existed instead^14^. We cleaned and normalized FAERS by removing duplicate individual case reports, mapping drug names to RxNorm concepts and outcomes to SNOMED-CT concepts based on a standardization method^16^. Finally, we mapped unique individual case reports with demographic information through ‘primaryid’ and ‘isr’.

In this study, we collected FAERS quarterly submissions from 2004 to the third quarter of 2018. The curated and standardized version consisted of 99,543,819 drug-ADE pairs. After removing missing values, and values out of 100 in age, a total of 75,748,043 pairs remained. We then divided patients into four age groups, 0-14 years old as children, 15-24 years old as youth, 25-64 years old as adult and *>*65 years old as senior on the basis of the criteria provided by World Population Prospects from The United Nations Department of Economic and Social Affairs^15^. In total, among these records, 2,408,437 pairs (3.18%) were from children, 2,648,358 pairs (3.50%) were from youth, 41,751,042 pairs (55.12%) were from adult and 28,940,206 pairs (38.20%) were from senior.

## Results and Analysis

After cleaning and standardization, there remained 4,580,113 unique drug-ADE pairs with 5,661 drugs and 19,198 ADEs in the data set. We applied the methodology to the data set. We removed pairs whose ADE is caused by the misuse of drugs, such as off label use and drug abuse. In the process, by screening drug labels, we also removed some possible false positive pairs caused by co-morbidities. For instance, the atorvastatin-coronary artery disease pair in adult might be spurious since atorvastatin is commonly used in the hyperlipidemia patients for the prevention of heart disease, which is a critical comorbidity. Finally, with confidence level at 0.999, we discovered 2,523 unique age-associated drug-ADE pairs and their highest age risks, among which 408 drugs and 775 adverse events were included. Full results are available in https://github.com/Zhizhen-Zhao/Age-Risk-Identification. Table 1 presents the number of drug-ADE pairs, drugs and ADEs in each age risk group.

**Table 1:** Number of drug-ADE pairs, drugs and ADEs in each age risk group

| Age Risk Group | Drug-ADE pairs | Drugs | Adverse Drug Events |
| --- | --- | --- | --- |
| Children | 629 | 208 | 342 |
| Youth | 672 | 226 | 284 |
| Adult | 255 | 94 | 143 |
| Senior | 531 | 175 | 244 |
| Children & Youth | 325 | 141 | 194 |
| Children & Adult | 7 | 4 | 7 |
| Children & Senior | 22 | 19 | 21 |
| Youth & Adult | 47 | 38 | 38 |
| Youth & Senior | 15 | 13 | 14 |
| Adult & Senior | 20 | 17 | 18 |

Among identified pairs, 1,966 (77.92%) pairs appeared in all age groups and 2,087 (82.72%) pairs have the single age risk group. Children and youth are the most susceptible groups among the whole population. Only 44 (1.74%) pairs show the highest risk in two age groups which are not continuous in classification. We identified the most prominent drug-ADE pairs that pose the highest risk to each single age group, the top 6 pairs are shown in Table 2. For the 10 most prescribed drugs in the U.S.^17^, we successfully identified atorvastatin, lisinopril, amlodipine, amoxicillin, omeprazole, losartan and metformin whose ADE pairs have significant age risk. Table 3 shows the most significant risks in a single age group from these drugs.

**Table 2:** Top 6 drug-ADE pairs within each higher age risk group

| Drug | Adverse Drug Event | ROR* | Adjusted P-Value |
| --- | --- | --- | --- |
| Children |  |  |  |
| risperidone | emotional disorder | 12.51 | 0 |
| rivaroxaban | gastrointestinal haemorrhage | 1.75 | 0 |
| loratadine | overdose | 27.25 | 0 |
| adalimumab | injection site pain | 2.29 | 4.82e-277 |
| warfarin | international normalised ratio increased | 1.88 | 3.82e-250 |
| risperidone | gynaecomastia | 8.68 | 4.38E-250 |
| Youth |  |  |  |
| misoprostol | haemorrhage | 5.01 | 7.15e-291 |
| finasteride | erectile dysfunction | 9.99 | 1.48e-179 |
| mifepristone | abortion incomplete | 1.98 | 3.88e-179 |
| alprazolam | cardiac arrest | 6.60 | 7.70e-166 |
| alprazolam | respiratory arrest | 7.20 | 2.29e-129 |
| doxycycline | haemorrhage | 5.75 | 2.43e-128 |
| Adult |  |  |  |
| aspirin | myocardial infarction | 3.06 | 1.63e-278 |
| isotretinoin | unintended pregnancy | 2.64 | 1.76e-248 |
| levonorgestrel | acne | 6.96 | 2.78e-217 |
| furosemide | cardiac failure congestive | 1.91 | 9.90e-213 |
| aspirin | cardiac failure congestive | 1.92 | 9.88e-167 |
| etonogestrel | pulmonary embolism | 2.46 | 1.99e-105 |
| Senior |  |  |  |
| vincristine | febrile neutropenia | 1.60 | 3.21e-256 |
| etanercept | rheumatoid arthritis | 1.51 | 9.51e-177 |
| salicylic acid | completed suicide | 2.74 | 1.19e-166 |
| cyclosporine | eye irritation | 6.78 | 1.30e-139 |
| ethanol | completed suicide | 2.55 | 2.37e-136 |
| medroxyprogesterone | breast cancer female | 1.99 | 3.51e-114 |

\* ROR is the odds ratio of the highest risk group over other groups.

**Table 3:** Top age risks posted by the most prescribed drugs in the US

| Drug | Adverse Drug Event | Age Risk Group | ROR* | Adjusted P-Value |
| --- | --- | --- | --- | --- |
| atorvastatin | type 2 diabetes mellitus | Children | 3.28 | 3.86e-10 |
|  | cardiogenic shock | Youth | 46.24 | 4.25e-12 |
|  | angina unstable | Adult | 1.93 | 2.46e-06 |
|  | atrial fibrillation | Senior | 4.91 | 1.17e-08 |
| lisinopril | renal impairment | Children | 4.76 | 5.74e-04 |
|  | shock | Youth | 16.03 | 5.88e-05 |
|  | coronary artery disease | Adult | 2.30 | 5.97e-09 |
| amlodipine | generalised oedema | Children | 12.32 | 5.06e-12 |
|  | shock | Youth | 11.30 | 1.85e-83 |
|  | myocardial infarction | Adult | 2.42 | 9.82e-20 |
| amoxicillin | urticaria | Children | 3.51 | 6.95e-08 |
|  | deep vein thrombosis | Youth | 4.49 | 6.95e-08 |
| omeprazole | injury | Youth | 12.06 | 4.5e-18 |
| metformin | glomerular filtration rate decreased | Children | 7.98 | 1.49e-06 |
|  | intentional overdose | Youth | 23.39 | 7.10e-40 |
|  | myocardial infarction | Adult | 1.35 | 3.71e-22 |
|  | decreased appetite | Senior | 1.89 | 2.72e-49 |

\* ROR is the odds ratio of the highest risk group over other groups.

In addition, we compared the distribution of significant age risks in each system organ class (SOC). All detected drug-ADE pairs are grouped at the SOC level. All 27 SOCs show the significant age risks, where ‘General disorders and administration site conditions’, ‘Injury, poisoning and procedural complications’ and ‘Vascular disorders’ are the top 3 classes that have age risks. Figure 2 presents top 15 SOCs that have all four single age risks. Adult risk is at a lower proportion for all classes. Seniors are at the highest risk for experiencing Infections and infestations, Renal and urinary disorders, Immune system disorders, Musculoskeletal and connective tissue disorders. Children are susceptible to Gastrointestinal disorders, Metabolism and nutrition disorders, Nervous system disorders, Investigations.

**Figure 2:**
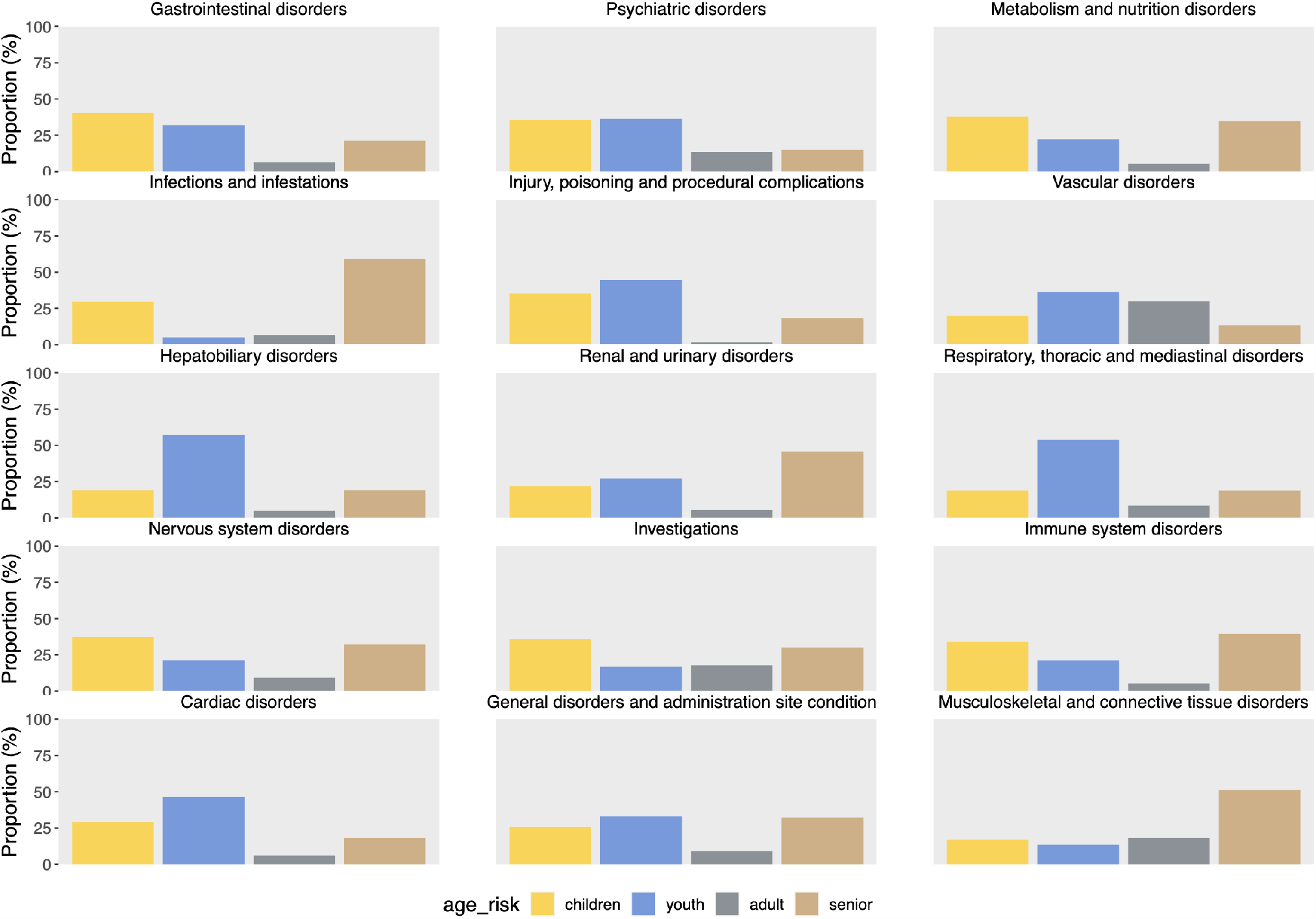
Proportion of single age risks in top 15 SOCs

### Case Study of Statin Drugs Induced ADEs in Children and Youth

In detected drug-ADE pairs with age risk, we filtered statin-related pairs that have age risk in either children or youth shown in Table 4. Our results well complemented current researches on statin-induced ADEs. The results indicated that the number of atorvastatin-related pairs with risk in children and youth seems to be significantly larger than other statin drugs, which demonstrates the variation of ADEs between different statin drugs. We also observed that statin-induced cardio and vascular events (bradycardia, cardiogenic shock, electrocardiogram qrs complex prolonged and hypertension) have a higher tendency in children and youth, which were not observed in previous clinical trials^18^. Meanwhile, a simvastatin-induced muscle symptom (myalgia) was found at a higher risk in the young population. Although this result is not consistent with a meta-analysis in 2008^19^which revealed that age (*>* 65 years old) is a risk factor for myopathy and rhabdomyolysis, almost all studies included in the meta-analysis were conducted in the adult population and they did not find enough data for the young population. In addition, we also conducted the Standardised MedDRA Queries (SMQs) evaluation regarding myopathy. The SMQ terms used in the analysis included Rhabdomyolysis/myopathy (SMQ code: 20000002) that consists of ten narrowly defined PT terms. We obtained a significant association (adjust p-value = 1.56e-06) between simvastatin and Statin associated muscle symptoms (SAMS) in the young population.

**Table 4:** Statin drugs-ADE pair with children or youth risks

| Statin Drug | Adverse Drug Event | Age Risk Group | Adjusted P-Value |
| --- | --- | --- | --- |
| atorvastatin | cardiogenic shock | Youth | 4.25e-12 |
|  | electrocardiogram qrs complex prolonged | Youth | 4.83e-12 |
|  | type 2 diabetes mellitus | Children | 3.86e-10 |
|  | squamous cell carcinoma of skin | Youth | 4.63e-10 |
|  | actinic keratosis | Youth | 1.81e-7 |
|  | hypertension | Youth | 1.73e-6 |
|  | seizure | Youth | 8.54e-5 |
|  | intentional overdose | Youth | 2.02e-4 |
|  | bradycardia | Youth | 2.69e-4 |
| lovastatin | completed suicide | Children | 2.50e-17 |
| rosuvastatin | completed suicide | Youth | 2.00e-11 |
| simvastatin | myalgia | Children | 4.53e-17 |
|  | personality change | Children | 1.32e-7 |
|  | completed suicide | Children & Youth | 3.26e-7 |

Our observation of the higher risk of children or youth in atorvastatin- and simvastatin-induced ADE could be attributed to dose. Current pediatric statin dosage recommendations are extrapolated from existing adult data^20^. Usual pediatric doses of atorvastatin and simvastatin for familial hypercholesterolemia (FH) patients are similar to those in adults (10–80 mg/day). Although some clinical trials in children proved atorvastatin and simvastatin’s efficacy and safety at these dose levels, there existed many limitations of these findings. The most critical one is the duration of statin therapy in these clinical trials. In clinical practice, patients with FH are subjected to continue with statin treatment for the rest of their lives once the therapy was initiated while the duration of trials could range from only 8 to 104 weeks. The accumulating dose of statin drugs could not be evaluated in the clinical trials.

On the other hand, pharmacogenetics might also cause a higher risk in children and youth. For instance, Wagner’s non-compartmental analysis of children and adolescent data^21^ demonstrated that each copy of the SLCO1B1 c.521C allele was associated with a 2.5-fold increase in simvastatin acid (SVA, the active form of simvastatin) systemic exposure, which was more pronounced than reported in adult studies.The 9-to the 10-fold range of AUC values noted within the c.521TT and c.521TC SLCO1B1 genotype groups exceeded the between-group variability, implying that additional factors may contribute to inter-individual variability in SVA systemic exposure in children and adolescents. This could induce a higher risk of simvastatin-induced myopathy in the children and youth population.

Thus, although the short- and intermediate-term efficacy and safety of statins have been confirmed by observational studies and meta-analyses^22^, there still exists some statin-induced ADEs that have not been fully explored due to biased study population or short evaluation period of drug effects. We found the variation of ADEs caused by different statin drugs and presented statins-induced ADEs that have the highest risk in children and youth. We explained the plausibility of results in terms of drug usage and pharmacogenetics. Our results could take the essential supplement in clinical trials, especially for the statin drugs-related safety issues.

## Conclusion

In this study, we developed a computational methodology to explore the differential risks of a drug causing an adverse drug event in different age groups. We performed multiple Chi-squared tests and disproportionality analysis to examine age risks in drug-ADE pairs. We also removed confounding effects due to age bias through logistic regression and likelihood ratio tests. Finally, We applied the methodology on FAERS data set and identified age-associated drug-ADE pairs as well as their highest age risk. FAERS data is subject to biases due to differential prescription and ADEs are usually dependent on physical conditions of the patients. Thus the proposed methodology can be further improved by adjusting possible confounders, such as gender,co-morbidities and co-prescribed drugs. Our results provided a new resource of age-related adverse events for drugs, which is important for the appropriate prescriptions for patients in different age groups. The new methodology could become an efficient tool in the improvement of precision medicine and drug safety supervision.

## Data Availability

All data produced in the present work are contained in the manuscript.

## Acknowledgement

This work was funded in part by the National Institute of General Medical Sciences (NIGMS) of NIH under award number R01GM141279.

## References

1. Aronson JK, Ferner RE. Clarification of terminology in drug safety. Drug Saf. 2005;28(10):851–70.

2. Chandak P, Tatonetti NP. AwareDX: Using machine learning to identify drugs posing increased risk of adverse reactions to women. SSRN Electron J. 2020.

3. Center for Drug Evaluation, Research. Preventable Adverse Drug Reactions: A Focus on drug interactions. http://Fda.gov. 2020. Available from: https://www.fda.gov/drugs/drug-interactions-labeling/preventable-adverse-drug-reactions-focus-drug-interactions

4. Considine J, Botti M. Who, when and where? Identification of patients at risk of an in-hospital adverse event: implications for nursing practice. Int J Nurs Pract. 2004;10(1):21–31.

5. Luo J, Eldredge C, Cho CC, Cisler RA. Population analysis of adverse events in different age groups using big clinical trials data. JMIR Med Inform. 2016;4(4):e30.

6. Huddleston JI, Wang Y, Uquillas C, Herndon JH, Maloney WJ. Age and obesity are risk factors for adverse events after total hip arthroplasty. Clin Orthop Relat Res. 2012;470(2):490–6.

7. Moore TJ, Weiss SR, Kaplan S, Blaisdell CJ. Reported adverse drug events in infants and children under 2 years of age. Pediatrics. 2002;110(5):e53.

8. Homma S, DiTullio MR, Sacco RL, Sciacca RR, Mohr JP, PICSS Investigators. Age as a determinant of adverse events in medically treated cryptogenic stroke patients with patent foramen ovale. Stroke. 2004;35(9):2145–9.

9. Pham M, Cheng F, Ramachandran K. A comparison study of algorithms to detect drug-adverse event associations: Frequentist, Bayesian, and machine-learning approaches. Drug Saf. 2019;42(6):743–50.

10. Loh W-Y, He X, Man M. A regression tree approach to identifying subgroups with differential treatment effects. Stat Med. 2015;34(11):1818–33.

11. Huang X, Sun Y, Trow P, Chatterjee S, Chakravartty A, Tian L, et al. Patient subgroup identification for clinical drug development. Stat Med. 2017;36(9):1414–28.

12. Seabroke S, Candore G, Juhlin K, Quarcoo N, Wisniewski A, Arani R, et al. Performance of stratified and sub-grouped disproportionality analyses in spontaneous databases. Drug Saf. 2016;39(4):355–64.

13. FDA Adverse Event Reporting System (FAERS). Available from: https://open.fda.gov/data/faers/

14. Yu Y, Chen J, Li D, Wang L, Wang W, Liu H. Systematic analysis of adverse event reports for sex differences in Adverse Drug Events. Sci Rep. 2016;6:24955.

15. World Population Prospects - Population Division - United Nations. http://Population.un.org. Available from: https://population.un.org/wpp/

16. Banda JM, Evans L, Vanguri RS, Tatonetti NP, Ryan PB, Shah NH. A curated and standardized adverse drug event resource to accelerate drug safety research. Sci Data. 2016;3(1):160026.

17. Paavola A. 10 most prescribed drugs in the US in Q1. Becker’s Hospital Review. Available from: https://www.beckershospitalreview.com/pharmacy/10-most-prescribed-drugs-in-the-u-s-in-q1.html

18. McCrindle BW, Ose L, Marais AD. Efficacy and safety of atorvastatin in children and adolescents with familial hypercholesterolemia or severe hyperlipidemia: a multicenter, randomized, placebo-controlled trial. J Pediatr. 2003;143(1):74–80.

19. Nguyen KA, Li L, Lu D, Yazdanparast A, Wang L, Kreutz RP, et al. A comprehensive review and meta-analysis of risk factors for statin-induced myopathy. Eur J Clin Pharmacol. 2018;74(9):1099–109.

20. Kearns GL, Abdel-Rahman SM, Alander SW, Blowey DL, Leeder JS, Kauffman RE. Developmental pharmacology—drug disposition, action, and therapy in infants and children. New England Journal of Medicine. 2003 Sep 18;349(12):1157–67.

21. Wagner JB, Abdel-Rahman S, Van Haandel L, Gaedigk A, Gaedigk R, Raghuveer G, et al. Impact of SLCO1B1 genotype on pediatric simvastatin acid pharmacokinetics. J Clin Pharmacol. 2018;58(6):823–33.

22. Avis HJ, Vissers MN, Stein EA, Wijburg FA, Trip MD, Kastelein JJP, et al. A systematic review and meta-analysis of statin therapy in children with familial hypercholesterolemia. Arterioscler Thromb Vasc Biol. 2007;27(8):1803–10.

